# Metformin reduces the risk of poor treatment outcomes among individuals with rifampicin-resistant tuberculosis and type-2 diabetes mellitus

**DOI:** 10.1101/2024.07.12.24310348

**Authors:** Argita D. Salindri, Mariam Gujabidze, Maia Kipiani, Nino Lomtadze, Nestani Tukvadze, Zaza Avaliani, Henry M. Blumberg, Hardy Kornfeld, Russell R. Kempker, Matthew J. Magee

## Abstract

We conducted a retrospective cohort study among individuals with rifampicin-resistant tuberculosis and diabetes to determine the association between metformin use and tuberculosis treatment outcomes. We found that individuals with metformin use had a significantly lower risk of poor tuberculosis treatment outcomes (adjusted RR=0.25, 95%CI 0.06 – 0.95) compared to those without.

## INTRODUCTION

It is well established that type-2 diabetes mellitus (diabetes) negatively impacts tuberculosis (TB); for instance, diabetes increases the risk of poor TB treatment outcomes including mortality during treatment. [1] The use of blood glucose lowering agents, such as metformin, may revert this adverse impact. [2] Metformin has been proposed as a potential candidate of adjunctive host-directed therapy for TB disease. [3, 4] However, there is a lack of clinical and epidemiological evidence to suggest whether metformin could improve TB treatment outcomes among individuals with drug-resistant TB (DRTB).

Limited evidence suggests that metformin may perform better in improving TB treatment outcomes compared to other diabetes medications. For example, a retrospective cohort study conducted using Taiwan’s health insurance database reported that individuals with TB and diabetes who were on metformin had lower risk of TB infection and disease progression compared those receiving sulfonylurea agents. [5] However, whether metformin is associated with improved TB clinical outcomes including sputum culture conversion remains poorly described. Furthermore, whether metformin is also associated with lower risk of post-TB mortality is also unknown. Thus, our study aimed to estimate the association between metformin use and different metrics of TB outcomes among individuals with DRTB and diabetes.

## METHODS

We conducted a retrospective cohort study among adult individuals (≥16 years) who were newly diagnosed with bacteriologically-confirmed pulmonary TB and had a record of pre-diabetes/diabetes indications (i.e., prior diabetes diagnosis or fasting blood glucose [FBG] levels of ≥100 mg/dL at TB treatment initiation). Individuals with bacterial strains resistant to rifampicin (i.e., rifampicin-resistant [RR]-TB) who started their treatment with second-line TB drugs at the National Center for Tuberculosis and Lung Diseases or two TB outpatient clinics located in Tbilisi, Georgia were eligible. Potentially eligible participants were pooled from Georgia’s TB surveillance system during the 2009 – 2017 period. In our analyses, diabetes status was determined by FBG ≥126.0 mg/dL at TB treatment initiation or a previous indication of diabetes diagnosis/treatment in medical charts. Pediatric cases, individuals with normoglycemic or pre-diabetes (i.e., FBG <126.0 mg/dL and no other diabetes indications), type-1 diabetes, clinical TB, or those treated outside of Tbilisi were excluded.

Our primary exposure was metformin use during TB treatment. Individuals with a record of metformin were assumed to take the medication as prescribed. Other study exposures included the use of other diabetes medications, diabetes history, years of living with diabetes, and diabetes complications. Our study outcomes included: a) time to sputum culture conversion, b) final TB treatment outcomes defined as “favorable” vs. “poor” according to the World Health Organizations’s guideline, [6] and c) all-cause mortality post-TB treatment. All-cause post-TB mortality was determined by cross-referencing individuals’ information (first and last name, date of birth, and unique national identifiers) with the death registry managed by the National Statistics Office of Georgia obtained on November 13^th^, 2019.

We used Cox proportional hazard models to estimate the association between metformin use and time to sputum culture conversion, as well as hazards of all-cause post-TB mortality. We used log-binomial logistic regression models to estimate the association between metformin use and poor TB treatment outcomes. Covariates included in the multivariable models were purposively selected due to the small sample size. All statistical analyses were performed using SAS version 9.4 (Cary, NC) with a p-value <0.05 considered statistically significant.

## RESULTS

During our study period, there were 1,416 adults with newly diagnosed RR-TB, 995 (70.3%) of which had available blood glucose information and 234 (16.5%) had either elevated blood glucose level at time of TB treatment initiation (i.e., FBG ≥100mg/dL) or a history of diabetes diagnosis/treatment. Of these, half (55.6%, 130/234) initiated treatment in Tbilisi; two (0.8%) of which had type-1 diabetes and 60 (25.6%) had FBG levels of 100-125mg/dL with no prior history of diabetes diagnosis/treatment (i.e., pre-diabetes), leaving 68 individuals with diabetes for analyses (Figure S1). The large majority of individuals with diabetes (60/68, 88.2%) had indications that they were on diabetes medications during TB treatment, but only 58 individuals had information on which diabetes medications were used; 31.0% (18/58) of which had a record of metformin use during TB treatment.

The median time to sputum culture conversion among 17 individuals who used metformin was 91 days (interquartile range [IQR] 61 – 122) vs. 121 days (IQR 91 – 187) among 33 who did not use metformin (p-value=0.89) (Table 1). After adjusting for age, gender and diabetes history, the hazard rate of sputum culture conversion among individuals who used metformin was 1.23 times (95% confidence interval [CI] 0.65 – 2.32) the hazard rate among those who did not use metformin (Table 2). The unadjusted risk of poor TB treatment outcomes was 11.8% among those who used metformin vs. 42.1% among those who did not use metformin (p-value=0.03). After adjusting for age, gender, and diabetes history, the risk of poor TB treatment outcomes remained significantly lower among individuals who used metformin compared to those who did not use metformin (adjusted risk ratio=0.25, 95%CI 0.06 – 0.95). Two individuals who used metformin died with the median time death of 31 months (IQR 2 – 60) post-TB treatment vs. 10 months (IQR 4 – 27) among five who did not use metformin and died post-TB (p-value=0.85). After adjusting for age, gender and diabetes history, the hazard rate of all-cause post-TB mortality among individuals who used metformin was 0.67 times (95%CI 0.10 – 4.49) the hazard rate among those who did not use metformin. The effect of other diabetes medications (i.e., sulfonylureas and insulin) on our study outcomes were provided in Table 2. We also provided the estimated effects of pre-diabetes on the three study outcomes in Table S1.

**Table 1.** Sputum culture conversion rate, risk of poor tuberculosis treatment outcomes, and all-cause post-tuberculosis mortality rate according to diabetes characteristics among individuals with rifampicin-resistant tuberculosis and diabetes, Georgia, 2009 – 2017 (N=68)

| Diabetes Characteristics | Total | TB Outcome Metrics |  |  |  |  |
| --- | --- | --- | --- | --- | --- | --- |
|  |  | Sputum culture conversion |  | Final TB treatment outcomes | All cause post-TB mortality |  |
|  |  | Converted/N (%) | Time to sputum culture conversion Median (IQR)* | Poor<sup>†</sup>/N (%) | Death/N (%) | Time to death Median (IQR)‡ |
| *Among those with known diabetes drugs prescription (N=58)* |  | **50/56 (89.3)** | **91 (61 – 121)** | **18/55 (32.7)** | **7/48 (14.6)** | **10 (2 – 53)** |
| Metformin use |  |  |  |  |  |  |
| No | 40 (69.0) | 33/38 (86.8) | 121 (91 – 187) | 16/38 (42.1) | 5/32 (15.6) | 10 (4 – 27) |
| Yes | 18 (31.0) | 17/18 (94.4) | 91 (61 – 122) | 2/17 (11.8) | 2/16 (12.5) | 31 (2 – 60) |
| Sulfonylureas use |  |  |  |  |  |  |
| No | 31 (53.5) | 27/30 (90.0) | 91 (61 – 122) | 10/30 (33.3) | 2/25 (8.0) | 19 (10 – 27) |
| Yes | 27 (46.5) | 23/26 (88.5) | 91 (61 – 97) | 8/25 (32.0) | 5/23 (21.7) | 4 (2 – 53) |
| Insulin use |  |  |  |  |  |  |
| No | 17 (29.3) | 16/17 (94.1) | 94 (81 – 115) | 3/17 (17.7) | 2/15 (13.3) | 29 (4 – 53) |
| Yes | 41 (70.7) | 34/39 (87.2) | 76 (61 – 121) | 15/38 (39.5) | 5/33 (15.2) | 10 (2 – 27) |
| Number of anti-diabetes medications |  |  |  |  |  |  |
| 1 |  |  |  |  |  |  |
| 2 or 3 | 35 (60.3) | 30/34 (88.2) | 91 (61 – 122) | 12/34 (35.3) | 4/29 (13.8) | 19 (7 – 40) |
|  | 23 (39.7) | 20/22 (90.9) | 93 (60 – 120) | 6/21 (28.6) | 3/19 (15.8) | 2 (1 – 60) |
| *Among all cohort (N=68)* |  | **58/66 (85.2)** | **91 (61 – 121)** | **23/65 (35.8)** | **8/56 (14.3)** | **19 (3 – 46)** |
| Diabetes history |  |  |  |  |  |  |
| Newly diagnosed diabetes | 20 (29.4) | 18/20 (90.0) | 91 (62 – 122) | 6/20 (30.0) | 4/16 (25.0) | 7 (3 – 25) |
| Known diabetes | 48 (70.6) | 40/46 (87.0) | 91 (61 – 120) | 17/45 (37.8) | 4/40 (10.0) | 40 (15 – 57) |
| Years living with diabetes |  |  |  |  |  |  |
| 0 – 5 years | 31 (45.6) | 27/29 (93.1) | 94 (62 – 121) | 9/29 (31.0) | 3/24 (12.5) | 10 (1 – 53) |
| 6 – 10 years | 14 (20.6) | 11/14 (78.6) | 62 (59 – 91) | 5/13 (38.5) | 2/14 (14.3) | 31 (2 – 60) |
|  |  | Sputum culture conversion |  | Final TB treatment outcomes | All cause post-TB mortality |  |
|  |  | Converted/N (%) | Time to sputum culture conversion Median (IQR)* | Poor<sup>†</sup>/N (%) | Death/N (%) | Time to death Median (IQR)<sup>‡</sup> |
| ≥11 years | 9 (13.2) | 7/9 (77.8) | 98 (33 – 153) | 4/9 (44.4) | 1/7 (14.3) | 27 (27 – 27) |
| *Unknown* | 14 (20.6) | 13/14 (92.9) | 89 (62 – 119) | 5/14 (35.7) | 2/11 (18.2) | 22 (4 – 39) |
| Diabetes complications |  |  |  |  |  |  |
| None | 34 (50.0) | 30/34 (88.2) | 63 (61 – 119) | 14/33 (42.4) | 5/33 (15.2) | 39 (10 – 53) |
| Any<sup>§</sup> | 23 (33.8) | 17/21 (81.0) | 95 (89 – 122) | 7/21 (33.3) | 3/16 (18.8) | 4 (1 – 27) |
| *Missing* | 11 (16.2) | 11/11 (100.0) | 94 (61 – 121) | 2/11 (18.2) | 0/3 (0.0) | NA |

\*Time to sputum culture conversion was defined as the time (measured in days) from TB treatment initiation to the first of two consecutive negative cultures that were at least 30 days apart
<sup>†</sup>Including 4 who failed the treatment, 18 who stopped the treatment (i.e., defaulted), and 1 who died during TB treatment (n=23)
<sup>‡</sup>Post-TB survival time (measured in months) was calculated from the end of TB treatment to the date of post-TB death or November 13<sup>th</sup>, 2019 for those who survived the follow-up period. One individual who died during TB treatment was excluded from the post-TB mortality analyses.
<sup>§</sup>Included a combinations of individuals with multiple diabetes complications. In our cohort, there were 6 individuals with neuropathy, 4 with nephropathy, 7 with retinopathy or other eye complications, 8 with heart disease, 1 with ascites, 2 with diabetic angiopathy, 1 with diabetic gangrene, and 1 with diabetic hepatitis
IQR – interquartile range; TB – tuberculosis
**Bold** indicates that the finding is statistically significant at $\alpha=0.05$

**Table 2.**
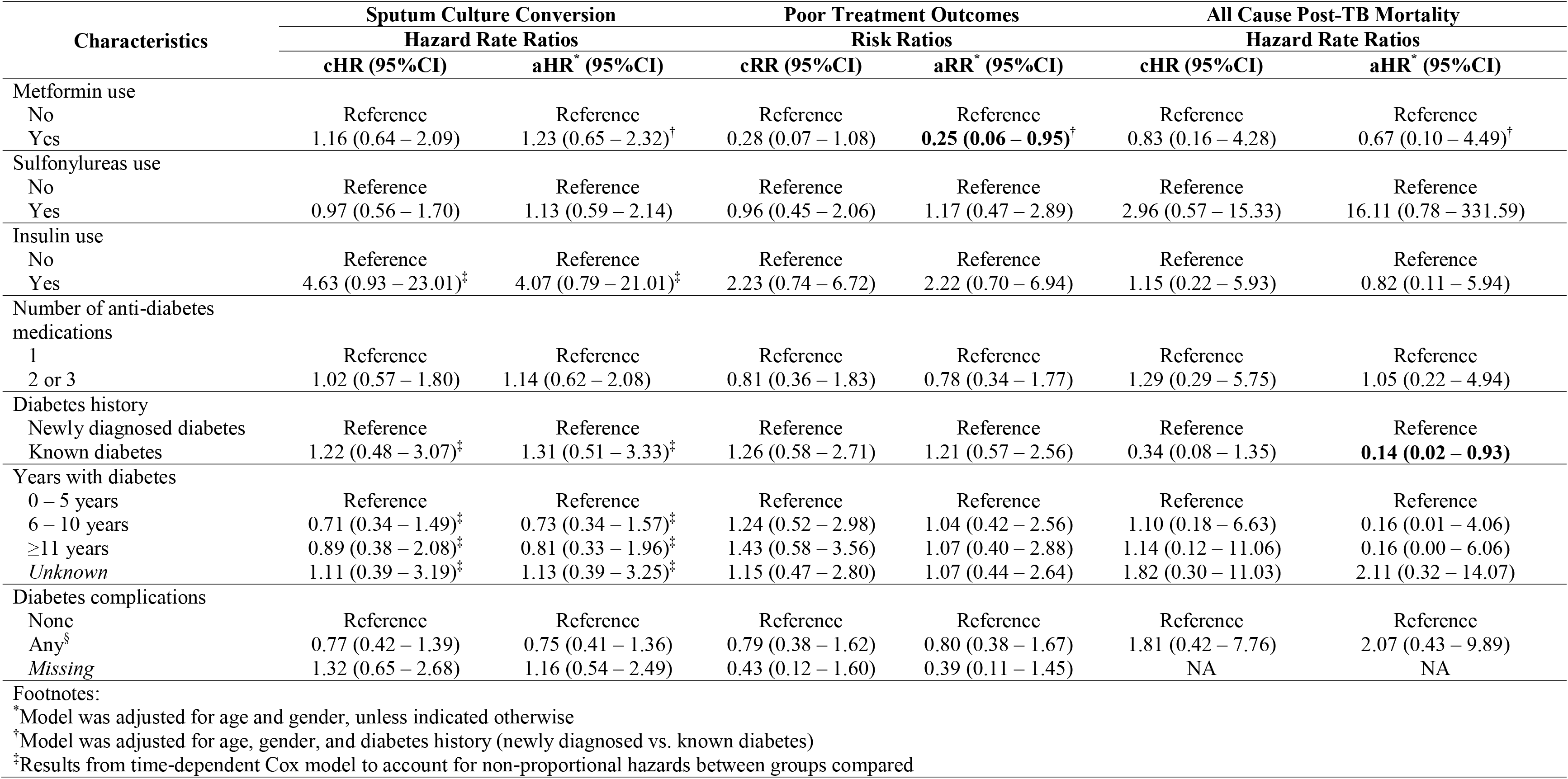
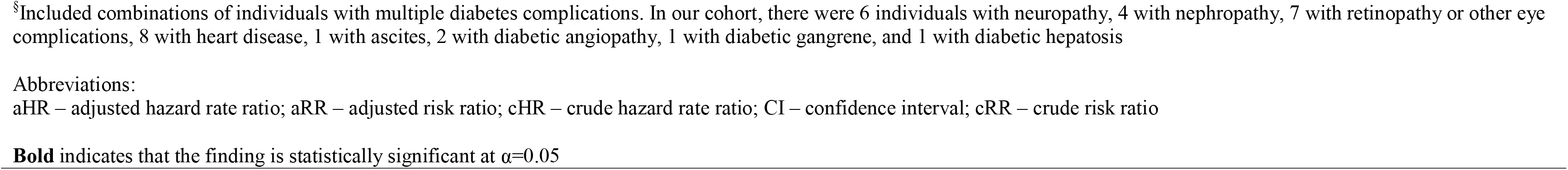
Crude and adjusted effects of metformin and other diabetes characteristics on a) sputum culture conversion, b) poor tuberculosis treatment outcomes, and c) all-cause post-tuberculosis mortality among individuals with rifampicin-resistant tuberculosis and diabetes, Georgia, 2009 – 2017 (N=68)

## DISCUSSION

We found that individuals with RR-TB and diabetes who used metformin had 75% lower relative risk of poor TB outcomes compared to individuals with RR-TB and diabetes who did not use metformin. Although non-sifinicant, individuals with a record of metformin use had higher rates of sputum culture conversion and lower rates of post-TB mortality compared to those without. Our finding on sputum culture conversion is consistent with a cross sectional study from South Korea which reported a modest difference in 2-month culture conversion among drug susceptible participants (61% metformin vs. 56% other diabetes medications, p=0.06). [7] Randomized studies assessing metformin’s mechanisms of action (including impact on blood glucose controls and reduction of inflammation) and comparing to other diabetes medications are needed.

Our cohort was limited by potential misclassification of metformin use and limited power to detect significant differences between diabetes medications. Metformin may have directly or indirectly reduced risk of poor outcomes, [8, 9] but it is also possible that metformin use in our cohort was related to less severe diabetes (e.g. fewer complications and less hyperglycemia) compared to those with insulin use. [10] We did not have sufficient measures to calculate metformin exposures during TB treatment and assess the dose-response relationship with our study outcomes. Larger prospective studies assessing adherence, duration of exposure of diabetes medications, and its effect on TB treatment ouctomes are still warranted.

Our study was conducted among a cohort of individuals with RR-TB and diabetes from Tbilisi, Georgia, a high RR-TB setting. Our study findings highlight the potiential role of metformin (compared to sulfonylureas or insulin) in reducing risk of poor outcomes during and after TB treatment among individuals with RR-TB and diabetes.

## Data Availability

Data that support the findings were collected for surveillance and treatment monitoring purposes; thus only accessible to investigators upon request to Georgia's National TB Program and Statistics Office. Individuals included in this cohort did not give written consent for their data to be shared publicly, and since the full data contain sensitive information including individuals' private health information (e.g., name and date of birth for matching purposes), the full dataset are not made publicly available. However, de-identified individual data, will be made available upon request to corresponding authors (ADS).

## ETHICS APPROVAL

This study was submitted to, reviewed, and approved by the Institutional Review Boards (IRBs) at Georgia State University and Emory University, Atlanta, USA and the ethics committee at National Center for Tuberculosis and Lung Diseases, Tbilisi, Georgia (FWA00020831). Waiver of informed consent was obtained from IRBs and ethics committee listed above as no human interactions occurred during the study process.

## AUTHOR CONTRIBUTIONS

ADS, MJM, and RRK conceived the study design. ADS, MG, and MK obtained the data. ADS performed the analyses. ADS and MJM interpreted the study results. ADS wrote the first draft of the manuscript and MJM revised the manuscript critically for important intellectual content. All authors reviewed, revised, and approved the final version of the manuscript.

## ACKNOWLEDGEMENTS

We would like to thank Mamuka Chincharauli for his assistance in pooling data from the Georgia’s TB online surveillance system.

## SUPPLEMENTAL MATERIALS

**Figure S1.**
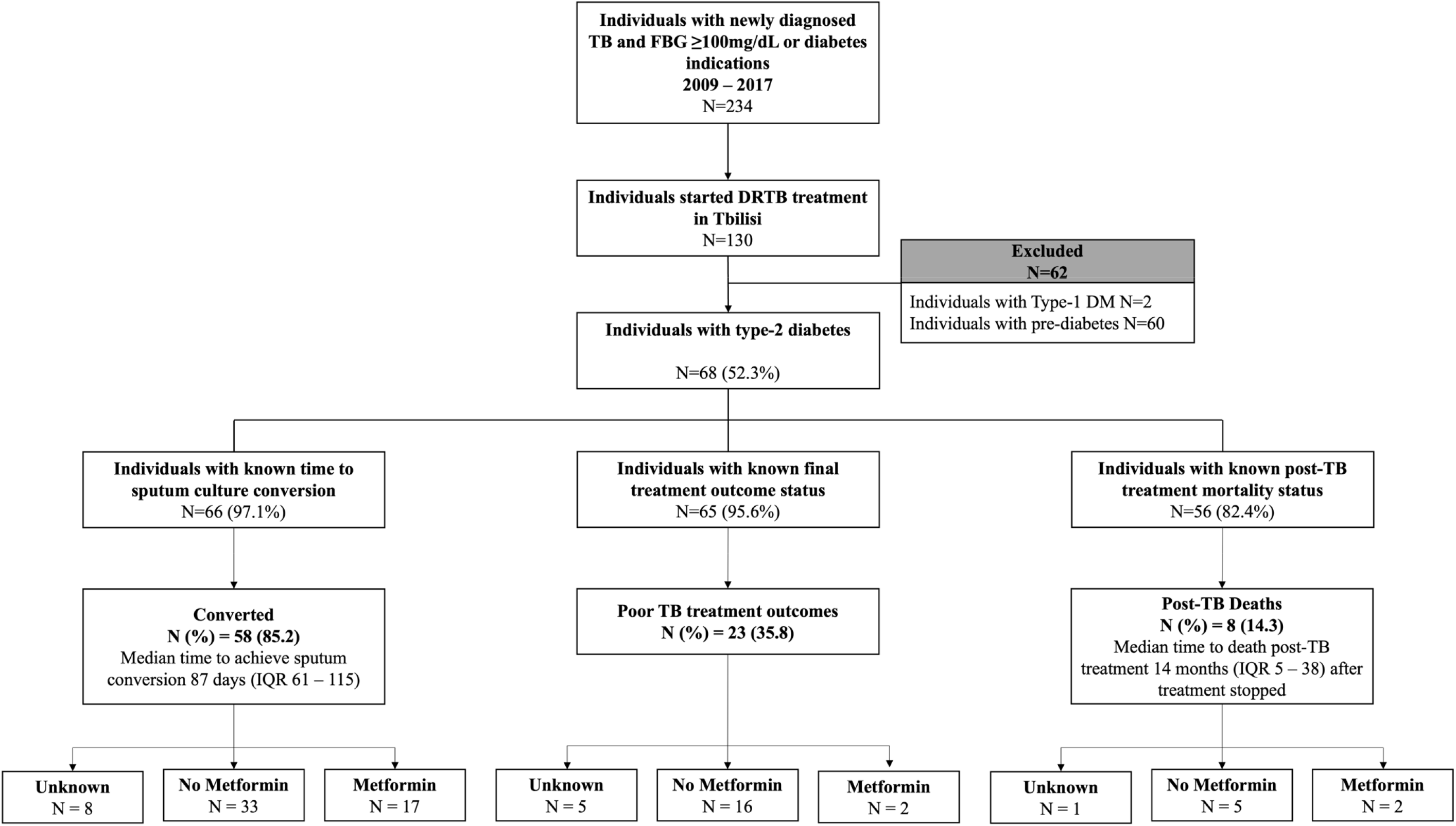
Study flow to depict the inclusion in study analyses.

**Table S1.** Clinical outcomes among individuals with rifampicin-resistant tuberculosis pre-diabetes and adjusted estimates compared to individuals with diabetes, Tbilisi, Georgia (N=128)

| Diabetes Status | Sputum Culture Conversion |  | Final TB Treatment Outcomes |  | Post-TB Mortality |  |
| --- | --- | --- | --- | --- | --- | --- |
|  | Converted | aHR* (95%CI) | Poor<sup>†</sup> | aRR* (95%CI) | Died | aHR* (95%CI) |
|  | N (%)=109 (85.2) |  | N (%)=44 (35.8) |  | N (%)=15 (13.9) |  |
|  | N (%) |  | N (%) |  | N (%) |  |
| Pre-diabetes<sup>‡</sup> | 51/56 (91.1) | 1.23 (0.79 – 1.92) | 21/58 (36.2) | 1.24 (0.74 – 2.09) | 5/48 (10.4) | 1.41 (0.42 – 4.78) |
| Diabetes | 58/66 (87.8) | Reference | 23/65 (35.4) | Reference | 8/56 (14.3) | Reference |

Abbreviations:
aHR – adjusted hazard rate ratio; aRR – adjusted risk ratio; CI – confidence interval;
\*Model was adjusted for age and gender
<sup>†</sup>Including 6 who failed the treatment, 36 who stopped the treatment (i.e., defaulted), and 2 who died during TB treatment
<sup>‡</sup>Fasting blood glucose 100 – 125.0 mg/dL with no other diabetes indications
**Bold** indicates that the finding is statistically significant at $\alpha=0.05$

## References

1. Huangfu, P., et al., The effects of diabetes on tuberculosis treatment outcomes: an updated systematic review and meta-analysis. Int J Tuberc Lung Dis, 2019. 23(7): p. 783–796.

2. Shewade, H.D., et al., Effect of glycemic control and type of diabetes treatment on unsuccessful TB treatment outcomes among people with TB-Diabetes: A systematic review. PLoS One, 2017. 12(10): p. e0186697.

3. Naicker, N., A. Sigal, and K. Naidoo, Metformin as Host-Directed Therapy for TB Treatment: Scoping Review. Frontiers in microbiology, 2020. 11: p. 435–435.

4. Yu, X., et al., Impact of metformin on the risk and treatment outcomes of tuberculosis in diabetics: a systematic review. BMC Infect Dis, 2019. 19(1): p. 859.

5. Pan, S.W., et al., The Risk of TB in Patients With Type 2 Diabetes Initiating Metformin vs Sulfonylurea Treatment. Chest, 2018. 153(6): p. 1347–1357.

6. WorldHealthOrganization, Definitions and Reporting Framework for Tuberculosis-2013 Revision. 2013, World Health Organization: Geneva.

7. Lee, Y.J., et al., The effect of metformin on culture conversion in tuberculosis patients with diabetes mellitus. Korean J Intern Med, 2018. 33(5): p. 933–940.

8. Yew, W.W., et al., Metformin as a host-directed therapeutic in tuberculosis: Is there a promise? Tuberculosis (Edinb), 2019. 115: p. 76–80.

9. Grzybowska, M., J. Bober, and M. Olszewska, [Metformin - mechanisms of action and use for the treatment of type 2 diabetes mellitus]. Postepy Hig Med Dosw (Online), 2011. 65: p. 277–85.

10. Costi, M., et al., Clinical characteristics of patients with type 2 diabetes mellitus at the time of insulin initiation: INSTIGATE observational study in Spain. Acta Diabetol, 2010. 47 Suppl 1(Suppl 1): p. 169–75.

